# Detection of maternal serum unmetabolized folic acid following multivitamin and mineral supplementation with or without folic acid after 12 weeks’ gestation: a randomized controlled trial

**DOI:** 10.1101/2022.07.18.22277791

**Authors:** Dian C Sulistyoningrum, Thomas R Sullivan, Monika Skubisz, Debra J Palmer, Simon Wood, Marten F Snel, Paul J Trim, Maria Makrides, Timothy J Green, Karen P Best

## Abstract

**Background:** Pregnant women are advised to take folic acid supplements before conception and during the first three months of pregnancy. Many women continue folic acid supplementation throughout pregnancy, and concerns have been raised about associations between excess folic acid intake and adverse child health outcomes. Unmetabolized folic acid (UMFA) is found in serum at higher folic acid intakes and has been proposed as a biomarker for excess folic acid intake.

**Objective:** To determine if removing folic acid from prenatal multivitamin and mineral supplements after 12 weeks of pregnancy reduces concentrations of serum UMFA at 36 weeks’ gestation.

**Design:** A double-blind, parallel-group, randomized controlled trial. Women with a singleton pregnancy 12-16-weeks’ gestation were randomly assigned to a multi-micronutrient supplement containing no folic acid (intervention) or 800 µg folic acid/day (control) from enrolment until 36 weeks’ gestation. Maternal serum was analyzed for UMFA and secondary outcomes of red blood cell and serum folate at 36 weeks’ gestation.

**Results:** UMFA was detected in most of the 103 randomized women (86% >limit of detection). However, only 12% (n=11/90) of serum samples were above the limit of quantification (0.55 nmol/L), preventing analysis of UMFA concentrations. Fewer women had detectable UMFA in the no folic acid group compared to the 800 µg folic acid group (72% [n=33/46] vs. 98% [n=43/44]; p = 0.001). Maternal serum and red blood cell folate concentrations were lower in the no folic acid intervention group compared to the control group (median 23.2 vs. 49.3 nmol/L, 1335 vs. 1914 nmol/L, respectively; p< 0.,001) and no woman was classified as folate deficient.

**Conclusions:** Removing folic acid from prenatal multivitamin and mineral supplements reduced the number of women with detectable UMFA at 36 weeks’ gestation, however, differences in UMFA concentration between treatment groups were not quantifiable.

## INTRODUCTION

Neural tube defects (NTDs) are serious birth defects caused by the failure of the neural tube to close properly, usually occurring around 28 days post-conception (1). Folic acid taken before conception and during early pregnancy is proven to reduce a woman’s risk of having an NTD-affected pregnancy (2-4); therefore, health authorities in most countries recommend women take a folic acid-containing supplement prior to conception (5, 6). In Australia, women are advised to take a supplement containing 500 µg of folic acid per day for a minimum of one month before trying to conceive and for the first three months of pregnancy (6). Although there is no conclusive evidence for any overall benefit of folic acid supplementation beyond 12 weeks gestation (31 trials involving 17,771 women) (7), many women continue to take folic acid supplements throughout their whole pregnancy, often at amounts up to 800 µg/day or higher (8). Because NTDs occur in the first month of pregnancy and many pregnancies are unplanned, more than 80 countries, including Australia, Canada, and the USA, have mandated folic acid fortification of staple foods with folic acid, further increasing folic acid exposure of pregnant women (9).

The common practice of continuing folic acid supplementation beyond the first trimester, especially in countries with folic acid fortification, is worrying due to increasing reports suggesting exposure to excess folic acid in late pregnancy may be associated with adverse child health outcomes, including an increased risk of allergic disease (10-12). Although findings from observational studies have been inconsistent, evidence from randomized controlled trials (RCTs) is lacking. The suggestion of risk necessitates further exploration of excess folic acid intake during pregnancy.

Folic acid is the synthetic form of folate not found naturally in food. Because of its high bioavailability and stability, it is the form of folate used in supplements and to fortify food (13). When consumed, folic acid is reduced and methylated to 5-methyltetrahydrofolate (5-MTHF) in the enterocyte or liver. At higher intakes, the enzymes required to convert folic acid to 5-MTHF are saturated, and the excess folic acid circulates in its unmetabolized form (UMFA) (14). UMFA has been proposed as a potential biomarker of excessive folic acid intake (14) and concerns have been raised over whether high concentrations of circulating UMFA may adversely affect the developing fetus (15). In acute dosing studies in non-pregnant individuals, UMFA rises rapidly after folic acid ingestion and falls over the following hours (14, 16). The greater the dose of folic acid, the higher the UMFA concentration and the longer it is detected in serum. The effect of chronic folic acid administration on UMFA is less clear.

UMFA has been detected in maternal blood samples in several population studies (17-20) and in one RCT in a country without mandatory fortification (21). However, there are no published RCTs investigating the effect of commonly used higher dose prenatal folic acid containing supplements, combined with background intake from mandatory fortification on UMFA concentration.

The importance of taking folic acid supplements in early pregnancy to reduce NTDs is not in doubt. However, supplementation beyond this time is in question. We aimed to investigate the effect of removing folic acid from prenatal supplements *after* 12 weeks’ gestation compared with the common practice of continuing folic acid supplementation of 800 µg/day throughout pregnancy on maternal serum UMFA at 36 weeks’ gestation.

## PARTICIPANTS AND METHODS

### Trial Design and Oversight

This trial was a multi-center, double-blind, placebo-controlled, parallel-group RCT. The trial protocol, published previously (22) was developed by the authors and approved by the Women’s and Children’s Health Network Research Ethics Committee – HREC/19/WCHN/018 and Flinders Medical Centre – SSA/20/SAC/61. The trial was conducted according to the 2007 National Statement on Ethical Conduct in Human Research and the Note for Guidance on Good Clinical Practice (CPMP/ICH/135/95) and prospectively registered with the Australia New Zealand Clinical Trials Registry - ACTRN12619001511123.

### Study Participants and Setting

Pregnant women living in South Australia were recruited to the trial between December 2019 and November 2020. Women with a singleton pregnancy between 12^+0^ and 16^+0^ weeks’ gestation, who were taking a folic acid-containing supplement and planning to continue it throughout pregnancy, were eligible to participate. Women were excluded if they; were carrying a fetus with a confirmed or suspected fetal abnormality, had a prior history of an NTD-affected pregnancy or were taking medications that interfere with folate metabolism. Women were recruited in person at their first antenatal clinic appointment or remotely through a Trial Recruitment Company (TrialFacts Australia, Melbourne, Victoria). They utilize an online digital marketing campaign and an electronic pre-screening survey.

### Randomization, Blinding, and Masking

After written informed consent was obtained, women were randomized by research personnel using a secure web-based randomization service and stratified by gestational age at trial entry 12^+0^ to ≤14^+0^ weeks’ or >14^+0^ to 16^+0^ weeks’ gestation. Allocation followed a computer-generated randomization schedule using randomly permuted blocks of sizes 4 and 6 (1:1 ratio), prepared by an independent statistician who was not involved with trial participants or data analysis. A unique and uninformative four-digit study identification number (Study ID) was assigned to each participant together with one of four colors for their group assignment (blue, pink, yellow, green). The intervention and control supplements were identical in size, shape, color, packaging, and labeling and identified by a colored label only. Participants, researchers, and laboratory personnel remained unaware of the group assignments until the data analysis was complete.

### Trial Interventions

Women in the intervention group received multivitamin and mineral supplements without folic acid. Women in the control group were assigned the same formulation with the addition of 800 µg of folic acid (Table S1 in the Supplementary Appendix). Following randomization, women were provided with two bottles, each containing 125 caplets, and advised to cease any folic acid-containing supplements for the duration of the trial. The assigned study supplements were taken once daily, orally from trial entry (12 weeks’ to 16 weeks’ gestation) until the day before the clinic visit and blood draw at 36 weeks’ gestation. Intervention and control supplements were formulated to provide daily multivitamin and mineral levels for prenatal supplementation and were manufactured in a licensed facility following the Code of Good Manufacturing Practice of Medicinal Products (23) by The Factors Group of Nutritional Companies Inc, (Coquitlam, British Columbia, Canada). The company had no other role in the trial.

### Data Collection

Baseline characteristics were collected at enrolment and included gestational age, maternal age, height and weight, race, education, pre-pregnancy and current supplement use, annual household income, parity and alcohol intake, and smoking in the three months leading up to pregnancy. Women were asked to complete an electronic 80-item food frequency questionnaire (FFQ) (The Dietary Questionnaire for Epidemiological Studies v3.2, Cancer Council, Victoria) at enrolment (baseline) and 34 weeks’ gestation to estimate folate intakes from foods. Adherence to the trial regimen and the occurrence of any adverse events were assessed by monthly electronic surveys sent by short message survey or phone call by study staff. Women returned for an in-person visit at 36 weeks’ gestation so that the number of unused caplets could be recorded, and a venous blood sample could be obtained by trained research personnel. Women were asked to refrain from taking their study supplement and consuming foods high in folic acid on the day of sample collection. Birth data including gestational age, birth weight, length, and head circumference were extracted from maternal and infant medical records or by parental report.

### 36 Week Sample Collection

A 10 ml non-fasting venous blood sample was collected into two evacuated containers containing no anticoagulant and ethylenediaminetetraacetic acid (EDTA) (BD Vacutainer®). The EDTA vacutainer was inverted 10 times and an aliquot was placed in a cryovial, diluted 1 in 11 with 1% ascorbic acid, and incubated for 30 minutes at 37°C. The serum vacutainer was left to clot at room temperature for at least 30 minutes. Vacutainers were centrifuged at 1500xg for 15 minutes at 4°C, serum and plasma were aliquoted into cryovials and stored at ^-^ 80°C until analyzed.

### Blood Analysis

A Complete Blood Count was performed using an automated hematology analyzer by SA Pathology (Adelaide, Australia). Serum UMFA was measured using the method of Hannisdal et al (24). Briefly, 10 µL (100 ng/mL) of d^4^ folic acid (F680302-0.5G, Novachem, Australia) was added to 100 µL of participant serum. Samples were deproteinized by adding 500 µL methanol, incubated at -20°C for 1 hour, and centrifuged for 15 minutes (12,000xg). The supernatant was collected and placed in a 96-well plate, dried under nitrogen at 65°C, and reconstituted with 100 µL 0.1% formic acid. Fetal bovine serum was used as blank to generate standard curve and quality control (QC) samples. Known concentrations of folic acid were spiked into fetal bovine serum to give the final concentrations of 0.25 limit of quantification (LOQ), 0.5, 1.0, 2.5, 5.5, 10, 20, 100 and 250 ng/mL for the standard curve and 0.625, 1.25, 7.5 and 50 ng/mL for the QC samples. Standards and QC samples were extracted using the same method as described above alongside the patient samples. Any standards or QCs outside +/-15% accuracy were excluded.

Liquid chromatographic separation of a 5 µL injection volume was achieved using an Acquity UPLC system (Waters Corporation, USA) fitted with a BEH Premier C18 (2.1 × 100 mm, 1.8 µm) chromatographic column maintained at 45°C. A linear gradient from 99% mobile phase A (0.1% formic acid) to 99% mobile phase B (acetonitrile 0.1% formic acid) over 2.5 minutes, followed by a 1 min hold at 99% B and a 1.5-minute re-equilibration at 99% A. Mass analysis was preformed using a 5500 Triple Quadrupole (Sciex, Canada) in Multiple Reaction Monitoring mode, transitions monitored are listed in Table S3 in the Supplementary Appendix. Data integration and analysis was preformed using Analyst 1.6.2 software (Sciex, Canada).

Whole blood and serum folate concentrations were determined using the microbiological method, using standardised kits from the US Centres for Disease Control and Prevention (US CDC; Atlanta, GA) (25-29). This method is based on the method of O’Broin and Kelleher (26), uses 96 well microplates, 5-methyl tetrahydrofolate (Merck Eprova) as a calibrator and chloramphenicol-resistant Lactobacillus rhamnosus (ATCC 27773TM) as the test organism. High and low quality controls (QC) provided by the US CDC whole blood and plasma folate, were run in quadruplets on every plate. RBC folate was calculated by subtracting plasma folate from whole blood folate and correcting for haematocrit.

As per instructions (27): if all QC results were within mean (2 SD) limits, the assay was accepted; if more than one of the QC results were outside of the mean (2 SD) limits or any of the QC results were outside of the mean (3 SD) limits, then the assay was rejected. Results from assay runs that passed QC were recorded only when the quadruplets were below 15%. If the coefficient of variation (CV) of the quadruplets was above 15%, the largest outlier was removed and the results recorded as long as the CV of the remaining triplicates was below 10%; otherwise, the sample measurement was repeated.

At the population level, WHO recommends RBC folate concentrations be >906 nmol/L in women of reproductive age to prevent NTDs. This RBC folate value was generated using folic acid as the calibrator (30, 31). We used a newer method recommended by the US CDC that uses 5-methyl tetrahydrofolate as the calibrator. Since 5-methyl tetrahydrofolate gives lower RBC folate concentrations than folic acid, we used a cutoff of >748 nmol/L to define the optimal RBC folate concentration for NTD risk reduction (31-33).

### Outcome Measures

The primary outcome was the concentration of UMFA in maternal serum at 36 weeks’ gestation. Secondary outcomes included maternal serum and RBC folate concentrations at 36 weeks’ gestation, and birth outcomes including: gestation age at birth, birth weight, birth length, and birth head circumference.

### Changes to Outcomes and Trial Design

We adapted some aspects of our methodology due to the COVID 19 pandemic. As per the recently released CONSERVE statement (34), we have described our original methods (22) and our adaptations as follows (35). When the trial commenced in December 2019 women were recruited from antenatal clinics and a baseline blood sample was collected at enrolment. In March 2020, due to COVID 19 restrictions in South Australia, in person enrolment was suspended and we could no longer collect the baseline blood sample. Screening methods were modified to include online screening coupled with a digital marketing campaign and e-consent using Research Electronic Data Capture (REDCap, Vanderbilt University). REDCap is a secure web application for building and managing online surveys and databases. Enrolment and all study visits up to 36 weeks’ gestation were conducted via telephone and supplements were couriered to participants. Birth data could no longer be extracted from medical records and were obtained by maternal report. Maternal and infant characteristics at birth were collected to establish comparison between groups as this study was not powered to evaluate clinical outcomes. We would caution about drawing conclusions due to the small sample size and lack of control for multiple testing.

### Sample Size and Statistical Analysis

A target sample of 90 women (45 per group) was chosen to provide >90% power to detect a standardized difference in mean UMFA concentration at 36 weeks’ gestation between groups of 0.60 (two-tailed alpha = 0.05, correlation between UMFA concentrations at baseline and 36 weeks’ gestation = 0.60) (21). Calculations were performed based on a standardized mean difference (mean difference divided by SD of outcome at 36 weeks’ gestation) due to considerable variability in the literature in the reported SD for UMFA concentration in pregnancy (10, 21).

All analyses were undertaken on an intention to treat basis according to a pre-specified statistical analysis plan (Supplementary Appendix). The trial was originally designed with serum UMFA concentration at 36 weeks (primary outcome) defined as a continuous outcome, with mean concentrations to be compared between groups using linear regression. However, a blinded review of 36-week UMFA concentrations unexpectedly revealed a high proportion of samples (88%) where the measurement was below the limit of quantification (LOQ, 0.55 nmol/L). Consequently, the primary outcome definition was changed to UMFA concentration at 36 weeks, dichotomized into above or below the limit of detection (LOD); this change was made before the statistical analysis plan was finalized and the treatment groups unblinded. The LOD was determined as a peak height of *ca* 4 times background noise. Its concentration was not calculated because it is not a true linear relationship to the standard curve at the LOD. The proportion of women with an UMFA concentration above the LOD was compared between groups using logistic regression. Secondary outcomes were analyzed using linear regression models, with log transformations applied where appropriate to better satisfy model assumptions. All analyses adjusted for gestational age at trial entry, since this was used to stratify the randomization, with analyses of birth anthropometrics additionally adjusted for infant sex. Statistical calculations were performed using Stata v16 (College Station, TX: StataCorp LP).

## RESULTS

### Trial Participants

A total of 103 women were randomized, 51 to the no folic acid group (intervention) and 52 to the 800 µg folic acid (control) group. After withdrawal of consent (n = 7), loss to follow-up (n = 4), unable to attend the clinic visit (n = 1) and preterm birth before 36 weeks’ gestation (n = 1), primary outcome data were available for 90/103 (87%) of women (**Figure 1**). The average age of women entering the trial was 31 years and more than 80% of the participants were Caucasian. The majority of women (87%) had completed secondary education and 55% had an annual household income higher than AUD$105,000. Overall mean total folate intake (±SD) was 585 ± 264 µg/day dietary folate equivalent (DFE) at baseline and 559 ± 253 µg/day DFE at 36 weeks. Folic acid added to food, and natural folate intake was 179 ± 120 µg/day, and 285 ± 108 µg/day at enrolment and 166 ± 116 µg/day, and 282 ± 104 at 36 weeks and did not differ markedly between the groups **(Table 1)**. Adherence to the trial supplements was similar between the intervention and control groups, with 85% of women who returned bottles (74%) consuming >80% of supplements to 36 weeks’ gestation, comparable with results from compliance questioning at study visits.

**Table 1.** Maternal baseline characteristics and folate intake at 36 weeks gestation^1^

| Characteristic | No folic acid<br>(n = 51) | 800 µg folic acid<br>(n = 52) |
| --- | --- | --- |
| Age, y | 30.7 ± 5.2 | 31.4 ± 4.4 |
| Gestational age at trial entry |  |  |
| 12 to < 14 wk | 32 (63) | 32 (62) |
| ≥14 to 16 wk | 19 (37) | 20 (38) |
| Maternal BMI at enrolment (n=94) | 25.2 ± 5.0 | 27.0 ± 6.2 |
| Ethnicity |  |  |
| European | 41 (80) | 44 (85) |
| Other | 10 (20) | 8 (15) |
| Completed secondary education | 46 (90) | 44 (85) |
| Annual household income |  |  |
| AUD\$70,000 or less | 9 (18) | 9 (17) |
| AUD\$70,001 - \$105,000 | 12 (24) | 7 (13) |
| AUD\$105,001 - \$205,000 | 23 (45) | 26 (50) |
| >AUD\$205,000 | 5 (10) | 7 (13) |
| Prefer not to disclose | 2 (4) | 3 (6) |
| Parity |  |  |
| 0 | 27 (53) | 20 (38) |
| Smoked tobacco in 3 mo before pregnancy | 5 (10) | 5 (10) |
| Consumed alcohol in 3 mo before pregnancy | 34 (67) | 41 (79) |
| Folic acid supplement intake at enrolment, µg/d |  |  |
| 250-<500 | 2 (4) | 1 (2) |
| 500-<750 | 24 (47) | 28 (56) |
| >750 | 25 (49) | 21 (42) |
| Folate intake at baseline, µg/d (n=88) |  |  |
| Total dietary folate<sup>2</sup> | 644 ± 298 | 528 ± 214 |
| Folic acid from fortified food | 204 ± 127 | 155 ± 110 |
| Natural food folate | 303 ± 124 | 268 ± 88 |
| Folate intake at 34 weeks, µg/d (n=84) |  |  |
| Total dietary folate<sup>2</sup> | 581 ± 269 | 538 ± 238 |
| Folic acid from fortified food | 179 ± 131 | 152 ± 100 |
| Natural food folate | 282 ± 105 | 281 ± 103 |

<sup>1</sup>Values are mean ± SD or n (%).<sup>2</sup>As Dietary Folate Equivalents = 1.7 x µg folic acid from fortified food + µg natural food folate

**Figure 1.**
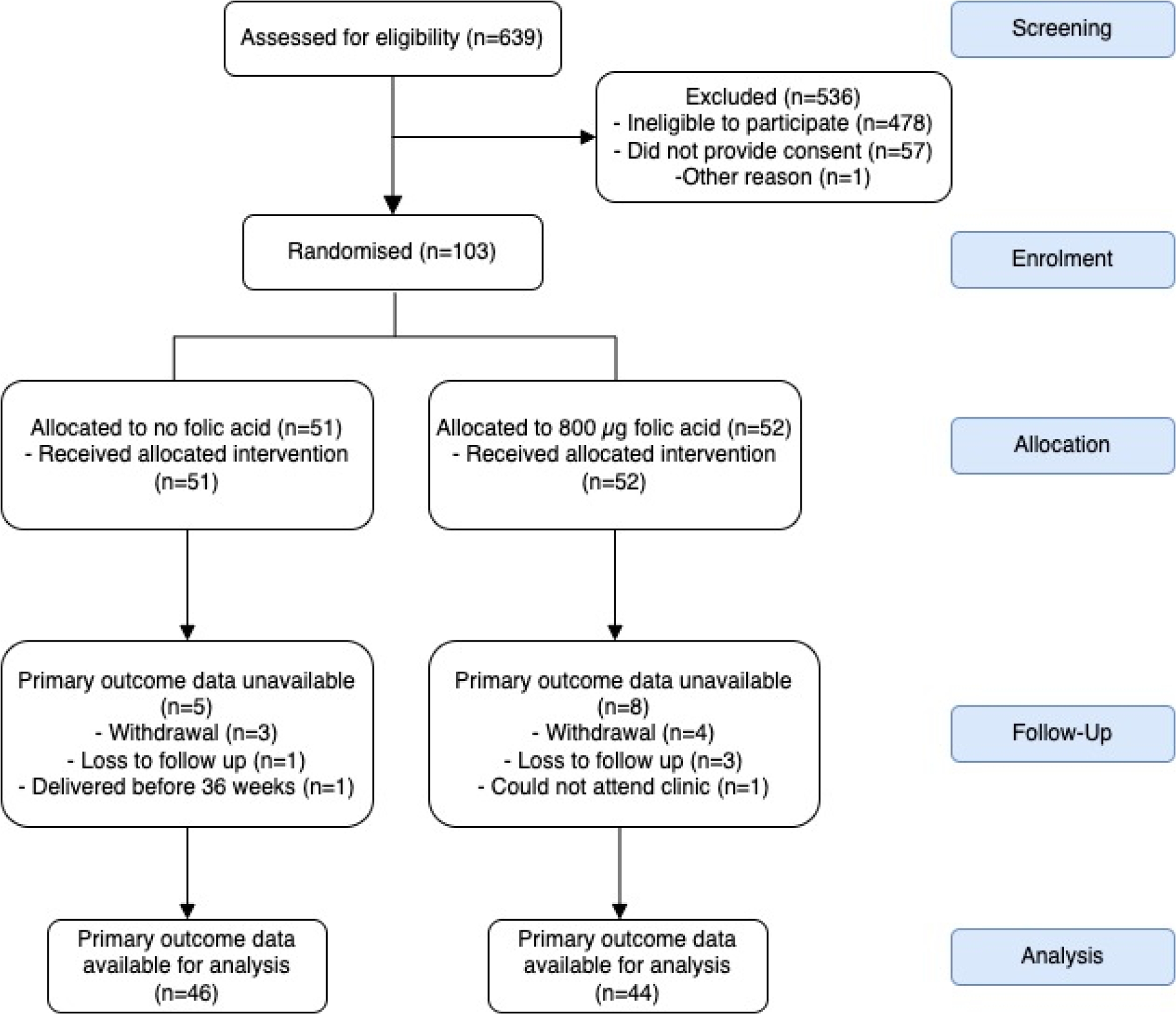
Participant Flow. NB. There were no protocol deviations relating to randomization errors.

### Outcomes

#### Unmetabolized Folic Acid (UMFA)

UMFA was detected in in 86% of samples of samples overall. UMFA ranged from undetectable to 14.2 nmol/L. Mean UMFA concentrations between the groups could not be reliably determined because only 12% (n=11/90) of sample were above the LOQ (0.55 nmol/L). However, the proportion of women with UMFA above the LOD at 36 weeks’ gestation was significantly lower in the no folic acid compared to the 800 µg folic group; 72% (n=33/46) vs. 98% (n=43/44), p=0.001. Of the women in the no folic acid group, 4/46 had UMFA concentrations above the LOQ compared with 7/44 in the 800 µg folic acid group which was not significantly different (χ^2^=1.09; p=0.47) (**Figure 2**).

**Figure 2.**
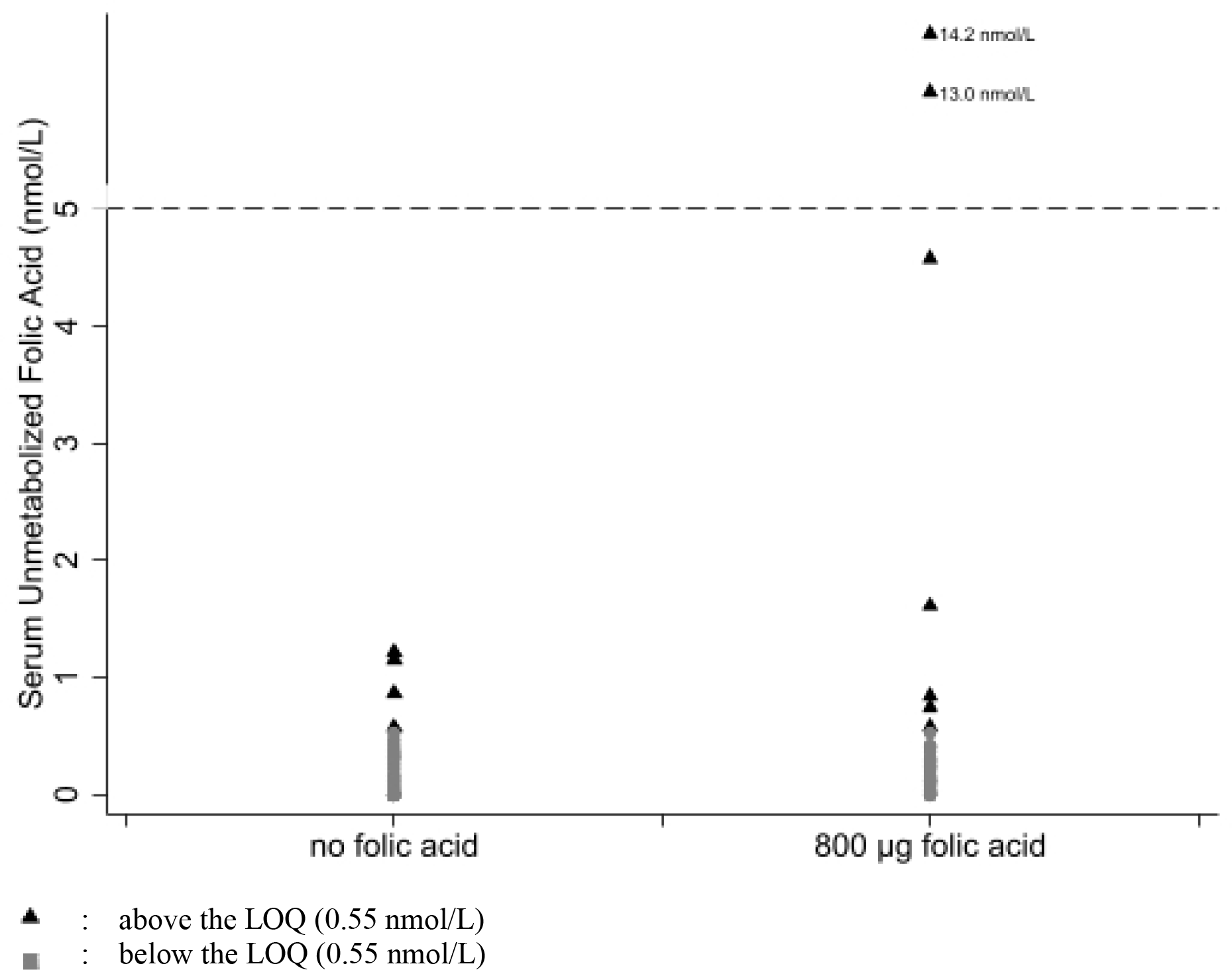
Serum unmetabolized folic acid in women at 36 weeks gestation.

#### Serum and Red Blood Cell Folate

Maternal serum folate concentrations were lower in the no folic acid group compared to the 800 µg folic acid group; median 23.2 nmol/L vs. 49.3 nmol/L, ratio of geometric means 0.56 (95% confidence interval (CI), 0.46 to 0.68 nmol/L), p<0.001 (**Table 2**). Similarly, median RBC folate concentrations were significantly lower in the no folic acid group than the 800 µg folic acid group; 1335 nmol/L vs. 1914 nmol/L, ratio of geometric means 0.69 (95% CI, 0.61 to 0.77), p<0.001 (Table 2). Serum and RBC folate concentrations were within normal clinical range in both the intervention and control groups.

**Table 2.** Blood folate concentrations at 36 weeks and neonatal outcome by treatment group

| Outcome | No folic acid<sup>1</sup> | 800 µg folic acid<sup>1</sup> | Treatment effect<sup>2</sup><br>(95% CI) | P-value |
| --- | --- | --- | --- | --- |
| Serum folate, nmol/L (n=90) | 23.2 (18.0, 28.4) | 49.3 (32.7, 57.7) | 0.56 (0.46, 0.68)<sup>3</sup> | < 0.001 |
| Red blood cell folate, nmol/L (n=90) | 1335 (1150, 1505) | 1914 (1534, 2300) | 0.69 (0.61, 0.77)<sup>3</sup> | < 0.001 |
| Gestational age at birth, wks (86) | 39.3 ± 1.7 | 39.0 ± 1.3 | 0.3 (-0.4, 1.0)<sup>4</sup> | 0.36 |
| Birth weight, g (n=90) | 3331 ± 519 | 3383 ± 473 | -44 (-255, 166)<sup>4</sup> | 0.68 |
| Birth length, cm (n=78) | 49.3 ± 2.8 | 49.9 ± 2.7 | -0.5 (-1.7, 0.8)<sup>4</sup> | 0.46 |
| Birth head circumference, cm (n=68) | 34.1 ± 2.1 | 35.0 ± 1.3 | -0.9 (-1.8, -0.1)<sup>4</sup> | 0.04 |

<sup>1</sup> Values are median (IQR) or mean ± SD
<sup>2</sup> Adjusted for gestational age at trial entry for all outcomes and additionally for infant sex for birth anthropometric outcomes
<sup>3</sup> Treatment effect expressed as a ratio of geometric means (95% CI)
<sup>4</sup> Treatment effect expressed as a mean difference (95% CI)

#### Birth Outcomes

There were no significant differences between the groups in birth outcomes, including gestational age, birth weight, and length, with the exception of head circumference which was lower in the no folic acid group compared to the 800 µg folic acid group (mean 34.1cm vs. 35.0 cm, difference = -0.9cm; 95% CI, -1.8 to -0.1), p=0.04 (Table 2).

#### Safety and adverse events

Adverse events were comparable between the groups with nausea the most common symptom overall at 1-week post-randomization (27%) and at 20 weeks’ gestation (29%) (Table S2 in the Supplementary Appendix). One infant in each group was admitted to the Neonatal Intensive Care Unit which was classified as a Serious Adverse Event. All serious adverse events were reviewed and categorized as unlikely related to the trial product or trial protocol.

## DISCUSSION

We examined the effect of removing folic acid from prenatal supplements after 12 weeks’ gestation on maternal UMFA in late pregnancy in a country with mandatory folic acid fortification. UMFA was detected in a smaller proportion of women randomized to the no folic acid supplement than the supplement containing 800 µg folic acid. Though UMFA was detected in the majority of women in our study (86%), most samples were below the LOQ (88% < LOQ 0.55 nmol/L) which meant we could not reliably report differences between groups in mean UMFA concentration.

Our results are similar to the findings of the only other published RCT investigating the effect of prenatal folic acid supplementation on UMFA concentration. Pentieva et al. reported that women randomized to folic acid supplements were more likely to have detectable plasma UMFA at 36 weeks’ gestation than women randomized to placebo (42% vs. 16%) (21) but also found no significant difference in mean concentration of UMFA between groups (0.13 ± SD 0.49 vs 0.44 ± SD 0.80, interaction p value = 0.38) (21). The dose of folic acid used by Pentieva et.al. (21) was lower than that found in common prenatal multivitamin and mineral supplements in Australia and many other countries, which range from 500 µg to 1,000 µg/day (20, 36). Furthermore, the Pentieva et al. study was conducted in Northern Ireland, which at the time only had voluntary folic acid fortification (21). We had expected to detect higher UMFA concentrations in our study as women in our control group were receiving double the amount of folic acid and were also exposed to a background of folic acid as a result of mandatory fortification in Australia. The prevalence of detectable UMFA in our population (86%) is similar to reports from observational studies in pregnant women in Australia (93%, > 0.03 to 244.7 nmol/L) (17), USA (81%, 0.23 to 1.47 nmol/L) (18) and Canada (97%, 0.00 to 0.91 nmol/L) (20). However, UMFA concentration differs substantially between studies and appears to be influenced by recent folic acid intakes which may explain the variability. Pfeiffer et al., reported detectable levels of UMFA in nearly all National Health and Nutrition Examination Survey (NHANES) participants (>95%, range >0.3 to 397 nmol/L) (37). NHANES is a representative sample of the US population and included men, women and children. Although 38% of NHANES survey participants were fasting >8 hours, Pfeiffer et al. reported that the detection of UMFA was evident regardless of fasting status, yet concentrations differed significantly by length of fasting (37).

We asked participants to avoid taking their study supplement on the day of their blood collection because we were interested in the chronic effect of folic acid supplementation on UMFA, not the acute effect as this is well established (14, 16, 38). Zheng et al. reported that following a single dose of 800 µg folic acid in 20 healthy male subjects, UMFA increased, peaking at around 2.5 hours in plasma but returned to undetectable levels within 12 hours (38). Although were able to detect UMFA in most participants, even those receiving no folic acid from study supplements (our intervention group), we had expected that chronic dosing of folic acid from early pregnancy would result in greater accumulation of UMFA. However, this was not the case and we were unable to quantify UMFA even in women who received 800 µg/d FA.

Comparison of our results with other studies of UMFA is difficult due to differences in measurement methods. We only quantified down to the LOQ, which means we did not report values below the lowest value in the standard curve for UMFA. Other researchers quantify down to the LOD, which if the term is being applied correctly, means that they are extrapolating below their lowest standard.

The differences we found in serum and RBC folate were as expected and consistent with other prenatal folic acid supplementation trials (21, 30, 39). At 36 weeks’ gestation, median serum folate was ∼26 nmol/L lower and median RBC folate was 600 nmol/L lower in the group receiving no folic acid versus 800 µg folic acid. Importantly all women remained above serum and RBC folate concentration indicative of deficiency, >6.8 nmol/L and >305 nmol/L, respectively (40).

Our study has many strengths, including a low attrition rate and high rate of supplement adherence. We asked women to refrain from taking their study supplement for 24 hours prior to their blood sample collection to reduce the variation in UMFA caused by recent high dose folic acid exposure. A limitation of our study is the absence of a baseline maternal blood sample at enrolment due to COVID-19 restrictions, which meant we were unable to examine changes in UMFA over time.

In conclusion, our trial showed that removing folic acid from prenatal multivitamin and mineral supplements reduced the number of women with detectable UMFA at 36 weeks’ gestation, however, differences in UMFA concentration between treatment groups were not quantifiable. The high within-subject variation when measured under standardized conditions and the lack of evidence in regard to what concentration of UMFA is normal, suggests that UMFA may not be the best biomarker to determine chronic excess folic acid exposure. There is no question that folic acid supplementation is essential, prior to, and in early pregnancy, but investigation of excess intake, especially in countries with mandatory fortication is warranted. High-quality randomized trials powered with clinical endpoints are needed to resolve concerns regarding the potential adverse effects of excess folic acid in late pregnancy on the health of children.

## Data Availability

Data described in the manuscript, code book, and analytic code will be made available upon request, pending submission of a proposal and subsequent approval by the Trial Steering Committee. Please forward requests to

## List of abbreviations

(CDC): Centre for Disease Control
(DFE): Dietary Folate Equivalent
(FFQ): Food Frequency Questionnaire
(LOD): Limit of Detection
(LOQ): Limit of Quantification
(NTDs): Neural Tube Defects
(RCT): Randomized Clinical Trial
(RBC): Red Blood Cell
(UMFA): Unmetabolized Folic Acid

## Acknowledgments

We would like to thank the participants who volunteered to be part of the trial and the antenatal staff at Women’s and Children’s Hospital and Flinders Medical Centre. We also would like to thank the clinical trial staff at South Australia Health and Medical Research Institute Women and Kids Theme. Study supplements (Prenuro®) were donated by Factors Group of Nutritional Companies Inc, Coquitlam, British Columbia, Canada.

## Statement of authors’ contributions to manuscript

KPB, TJG, DCS, MM, DJP and MS conceived the trial and proposed the original trial design; DCS programmed the study data management system, conducted analysis of blood samples for serum and blood cell folate, and participated in data collection; TRS advised on sample size calculations, trial design, data management and conducted the statistical analysis; PJT and MFS developed and validated the method for unmetabolized folic acid and DCS prepared the samples for UMFA analysis; SW and TJG designed the prenatal supplement and SW coordinated the manufacture of the supplements. DCS, TJG and KPB interpreted the data and drafted the manuscript; all authors provided critical input into the manuscript and all approved the final version. KPB was responsible for study oversight, data quality and the day to day management of the trial.

## Supplementary Appendix

This appendix has been provided by the authors to give readers additional information about their work.

### Supplement to

**Supplementary Table S1.** Ingredients of Supplements for Intervention (no folic acid) and Control (800 µg folic acid) Groups

| Ingredients | No folic acid | 800 µg folic acid | unit |
| --- | --- | --- | --- |
| folic acid | 0 | 0.8 | mg |
| calcium | 250 | 250 | mg |
| Iron | 27 | 27 | mg |
| thiamine | 1.4 | 1.4 | mg |
| riboflavin | 1.4 | 1.4 | mg |
| niacinamide | 18 | 18 | mg |
| vitamin B-6 | 1.9 | 1.9 | mg |
| vitamin B-12 | 2.6 | 2.6 | mcg |
| pantothenic acid | 6 | 6 | mg |
| biotin | 30 | 30 | mg |
| vitamin C | 85 | 85 | mg |
| vitamin E | 13.5 | 13.5 | IU |
| magnesium | 50 | 50 | mg |
| zinc | 7.5 | 7.5 | mg |
| manganese | 2.0 | 2.0 | mg |
| iodine | 0.22 | 0.22 | mg |
| copper | 1 | 1 | mg |
| selenium | 30 | 30 | mcg |
| Vitamin D3 | 10 | 10 | mcg |
| b-carotene | 2500 | 2500 | IU |

**Supplementary Table S2.** Adverse Events

|  | No folic acid | 800 µg folic acid | Fisher exact test p-value |
| --- | --- | --- | --- |
| **1-week post randomization** |  |  |  |
| Diarrhea | 2/50 (4%) | 3/49 (6%) | 0.68 |
| Nausea | 13/51 (25%) | 14/49 (29%) | 0.82 |
| Vomiting | 7/51 (14%) | 6/49 (12%) | >0.99 |
| Eructation | 3/51 (6%) | 5/48 (10%) | 0.48 |
| Constipation | 6/51 (12%) | 7/49 (14%) | 0.77 |
| Other symptom | 2/51 (4%) | 2/49 (4%) | 1.00 |
| **20 weeks gestation** |  |  |  |
| Diarrhea | 4/47 (9%) | 6/46 (13%) | 0.52 |
| Nausea | 13/47 (28%) | 14/46 (30%) | 0.82 |
| Vomiting | 7/47 (15%) | 6/46 (13%) | >0.99 |
| Eructation | 10/47 (21%) | 11/46 (24%) | 0.81 |
| Constipation | 15/47 (32%) | 19/46 (41%) | 0.39 |
| Other symptom | 1/47 (2%) | 6/46 (13%) | 0.06 |
| **24 weeks gestation** |  |  |  |
| Diarrhea | 2/45 (4%) | 3/43 (7%) | 0.67 |
| Nausea | 4/45 (9%) | 10/43 (23%) | 0.08 |
| Vomiting | 1/45 (2%) | 4/43 (9%) | 0.20 |
| Eructation | 7/45 (16%) | 11/43 (26%) | 0.30 |
| Constipation | 11/45 (24%) | 12/43 (28%) | 0.81 |
| Other symptom | 2/45 (4%) | 2/43 (5%) | >0.99 |
| **28 weeks gestation** |  |  |  |
| Diarrhea | 4/46 (9%) | 7/43 (16%) | 0.34 |
| Nausea | 3/46 (7%) | 5/43 (12%) | 0.48 |
| Vomiting | 0/46 (0%) | 1/43 (2%) | 0.48 |
| Eructation | 6/46 (13%) | 8/43 (19%) | 0.57 |
| Constipation | 8/46 (17%) | 16/43 (37%) | 0.06 |
| Other symptom | 1/46 (2%) | 3/43 (7%) | 0.35 |
| **32 weeks gestation** |  |  |  |
| Diarrhea | 4/46 (9%) | 3/44 (7%) | >0.99 |
| Nausea | 2/46 (4%) | 7/44 (16%) | 0.09 |
| Vomiting | 1/46 (2%) | 2/44 (5%) | 0.61 |
| Eructation | 4/46 (9%) | 8/44 (18%) | 0.23 |
| Constipation | 7/46 (15%) | 9/44 (20%) | 0.59 |
| Other symptom | 1/46 (2%) | 2/43 (5%) | 0.61 |
| **36 weeks gestation** |  |  |  |
| Diarrhea | 3/45 (7%) | 4/43 (9%) | 0.71 |
| Nausea | 5/45 (11%) | 6/43 (14%) | 0.76 |
| Vomiting | 1/45 (2%) | 2/43 (5%) | 0.61 |
| Eructation | 2/45 (4%) | 5/43 (12%) | 0.26 |
| Constipation | 6/45 (13%) | 7/43 (16%) | 0.77 |
| Other symptom | 4/45 (9%) | 2/43 (5%) | 0.68 |

**Supplementary Table S3.**
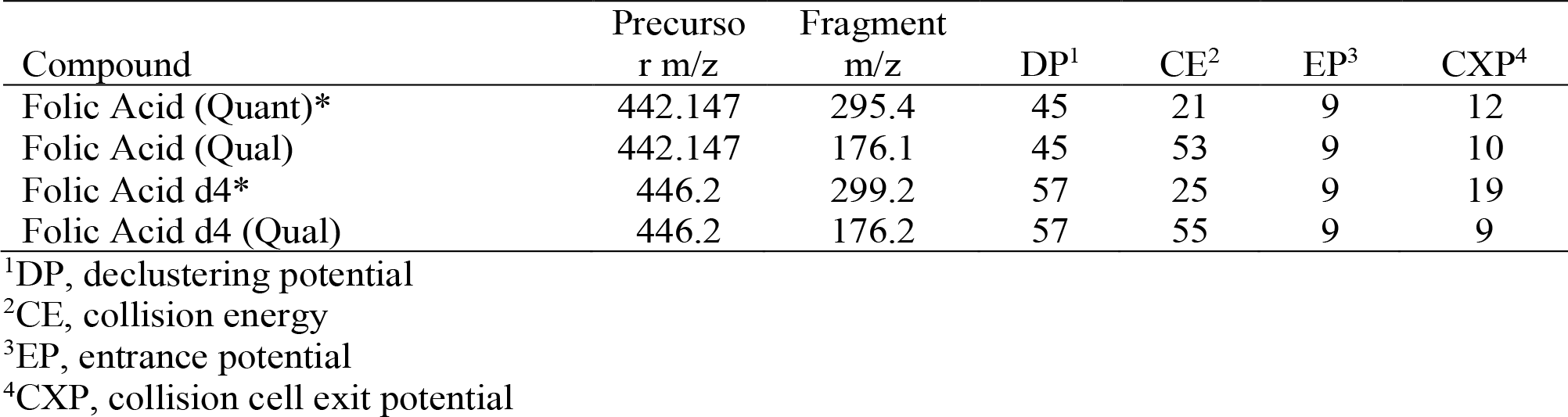
Transitions used for unmetabolized folic acid quantification

